# Standard pleural interventions are not high-risk aerosol generating procedures

**DOI:** 10.1101/2021.04.12.21255307

**Authors:** DT Arnold, FKA Gregson, S Sheikh, FW Hamilton, H Welch, A Dipper, GW Nava, AERATOR group, JW Dodd, AO Clive, BR Bzdek, JP Reid, NA Maskell

**Affiliations:** Academic Respiratory Unit, University of Bristol, BS10 5NB; Bristol Aerosol Research Centre, School of Chemistry, University of Bristol, BS8 1TS; MRC Integrative Epidemiology Unit, University of Bristol, BS8 2HW; Infection Sciences, North Bristol NHS Trust, BS10 5NB

**Author notes:** Corresponding author: Dr David Arnold, NIHR Doctoral Research Fellow, Academic Respiratory Unit, University of Bristol, Level 2 Learning & Research Building, Southmead Hospital, Bristol BS10 5NB, UK. AERATOR group (in alphabetical order): The AERATOR group consists of (in alphabetical order): Arnold, D; Brown, J; Bzdek, B; Davidson, A; Dodd, JW; Gormley M; Gregson, F; Hamilton, F; Maskell, N; Murray, J; Keller, J; Pickering, A.E; Reid, J; Sheikh, S; Shrimpton, A.

**Keywords:** COVID-19, Infection Control, Pleural Disease

## Abstract

No evidence exists regarding the risk of aerosolisation from pleural procedures. This study used two discrete methodologies, in an environment with no background aerosol interference, to measure aerosol generation from 10 different pleural procedures (3 medical thoracoscopies, 3 indwelling pleural catheter insertions, 1 therapeutic thoracentesis, and 3 indwelling pleural catheter removals). The measurements indicated that, any aerosol production during these procedures was significantly lower than aerosols produced by the patient breathing or coughing. Pleural procedures should not be considered aerosol generating. We hope this study informs future iterations of guidelines on the appropriate use of PPE when performing these procedures.

## Introduction

The coronavirus epidemic has focused attention on the risk of aerosol generating procedures (AGPs) in healthcare. SARS-CoV-2 has been isolated from pleural fluid which has the potential to infect staff if viraemic fluid were aerosolised during procedures^(1, 2)^. However, evidence for aerosol generation from pleural procedures is very limited. Current guidelines for appropriate use of personal protective equipment (PPE) while performing pleural procedures are based on expert opinion and application of the precautionary principle^(3)^. We set out to quantify the aerosol generation from pleural procedures compared to aerosol sampled during normal respiratory activities of breathing and coughing.

## Methods

This study performed as part of the AERATOR study assessing the risk of aerosolised transmission of SARS-CoV-2 in healthcare. Ethical approval was granted by North-West Research Ethics Committee (Ref:20/NW/0393).

Aerosol measurements were recorded simultaneously using two devices: an Optical Particle Sizer (OPS, TSI Inc.model 3330, USA, sampling flow rate 1L.min-1, samples 0.3-10µm diameter particles with a sampling period set as 1s) and an Aerodynamic Particle Sizer (APS, TSI Inc.model 3330, USA, sampling flow rate 1L.min-1, sheath flow 4L.min-1, samples 0.5-20µm diameter particles with a sampling period set as 1s). Technical specifications detailed in a previous publication (https://www.medrxiv.org/content/10.1101/2021.01.29.21250552v1), with aerosol sampled through a funnel 10cm from the operating site. The proximity of the sampling location is to ensure the source strength is measured without the convolved effects of dispersion and dilution at further distance from the source.

To reduce background aerosol concentration all procedures were performed in an ultra-clean laminar flow operating theatre (EXFLOW 32, Howarth Air Technology, UK) with high efficiency particulate air (HEPA) filtration and air supply rate of 1200m^3^.s^-1^ (550-650 air changes/hr). The air flow is 0.2m.s^-1^ at 1m above the floor below the laminar flow. To demonstrate that this airflow does not affect the sampling efficiency of aerosol generated under the laminar flow, the aerosol generated by voluntary coughing and breathing was sampled in the same position, 10 cm from a subject’s face.

Given the different pleural procedures have common themes, the procedures were sub-classified into 5 different elements (see Table 1). We also measured any aerosol produced from 3 pleural fluid management systems.

**Table 1:** The peak concentration sampled during a particular procedure of activity in this study (that reported in Fig. 1) and the mean particle concentration sampled throughout, for both the APS and OPS instruments.

|  | *n* |  | Peak concentration / cm-3 |  | Mean concentration / cm-3 |  |
| --- | --- | --- | --- | --- | --- | --- |
|  | APS | OPS | APS | OPS | APS | OPS |
| Pleural Anaesthesia | 8 | 10 | 0.023 | 0.036 | $9.52 \times 10^{-4}$ | $1.44 \times 10^{-3}$ |
| Therapeutic Thoracentesis | 3 | 5 | 0.04 | 0.036 | $1.10 \times 10^{-3}$ | $9.66 \times 10^{-4}$ |
| Chest drain insertion | 5 | 6 | 0.048 | 0.04 | $9.31 \times 10^{-4}$ | $1.04 \times 10^{-3}$ |
| Open pleural space at thoracoscopy | 2 | 3 | 0.03 | 0.04 | $1.19 \times 10^{-4}$ | $1.09 \times 10^{-4}$ |
| Chest drain removal | 3 | 3 | 0.1 | 0.018 | $7.89 \times 10^{-3}$ | 0.0124 |
| IPC Aspiration | 4 | 4 | 0.015 | 0.03 | $4.02 \times 10^{-4}$ | $5.07 \times 10^{-4}$ |
| Underwater seal bottle | 3 | 4 | 0.04 | 0.03 | $6.56 \times 10^{-4}$ | $9.84 \times 10^{-4}$ |
| Thopaz device | 2 | 2 | 0.06 | 0.09 | $1.59 \times 10^{-3}$ | $2.50 \times 10^{-3}$ |
| Background | 10 | 12 | 0.078 | 0.078 | $1.05 \times 10^{-3}$ | $1.37 \times 10^{-3}$ |
| Cough | 4 | 5 | 1.95 | 2.512 | - | - |
| Breathing | 5 | 6 | - | - | 0.276 | 0.223 |

## Results

Ten patients requiring pleural procedures (3 medical thoracoscopies, 3 indwelling pleural catheter insertions (15.5Fr), 1 therapeutic aspiration (6Fr), 3 indwelling pleural catheter removals)) were recruited to the study, with further 2 further patients with chest tubes already in-situ for pneumothorax with ongoing air leak. The majority of patients were male (10/12) with a median age of 76 (IQR 72-79).

Figure 1 (logarithmic Y axis) shows the peak aerosol number concentration sampled during each procedure compared to peak number concentrations from coughing and mean number concentrations from breathing. For most procedures, the peak number concentration was of similar magnitude to or less than the mean aerosol number concentration measured during breathing from the same patients or from healthy volunteers from a previous study (https://www.medrxiv.org/content/10.1101/2021.01.29.21250552v1) and was significantly less than the peak number concentration detected from a cough. The mean concentration for all procedures is reported in Table 1 and is typically much less (up to two orders of magnitude) smaller than the mean concentration sampled when a subject is breathing. Again, it should be stressed that breathing is a sustained activity while coughs and these clinical interventions lead to transient events.

**Figure 1:**
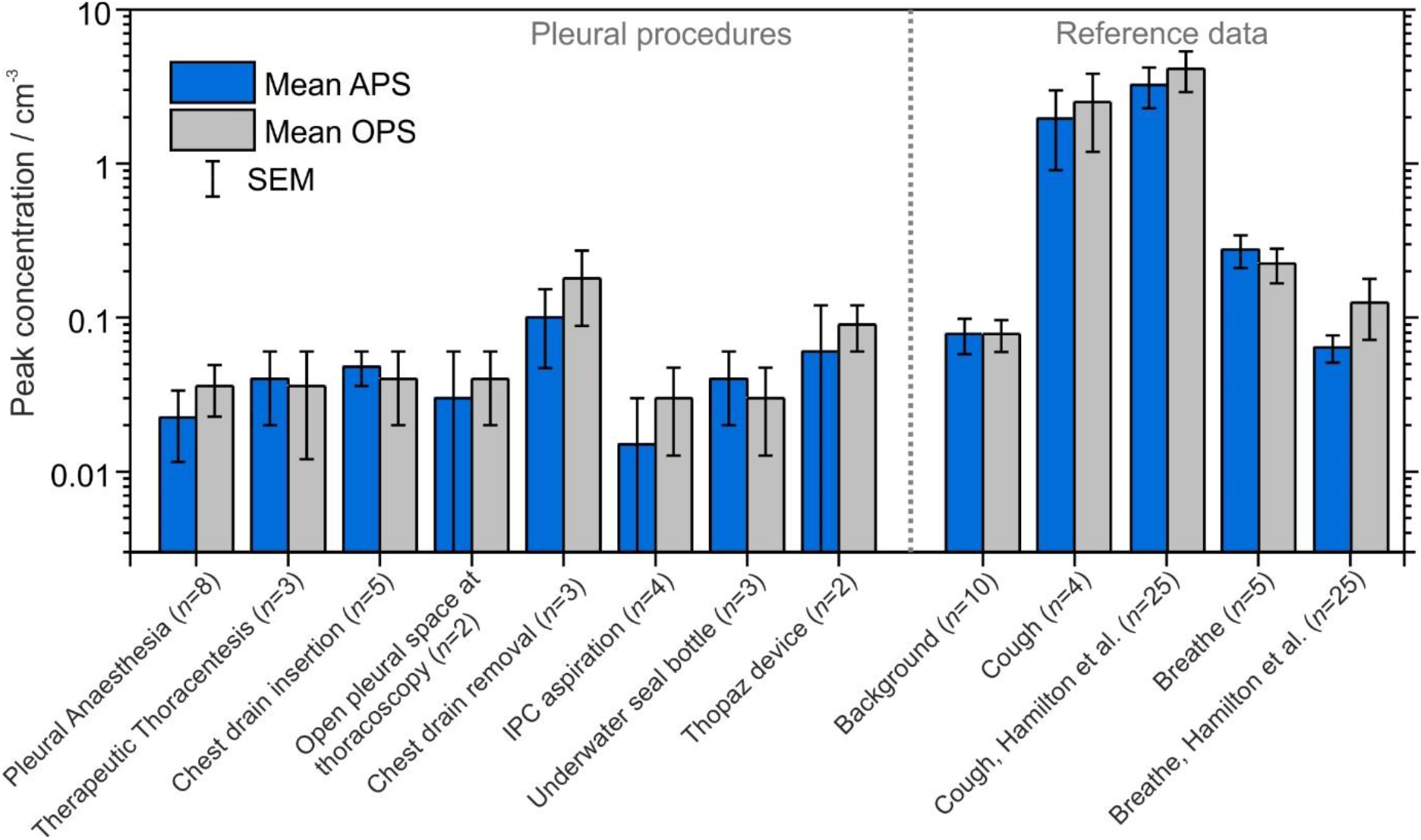
Bar chart showing the peak aerosol number concentration sampled by the APS and OPS methods during different procedural elements compared to mean aerosol number concentrations measured during breathing and peak aerosol number concentrations measured during cough.

Figure 2 illustrates the difference between the particle number concentration sampled during a single procedure compared to breathing or coughing for a patient undergoing a medical thoracoscopy.

**Figure 2:**
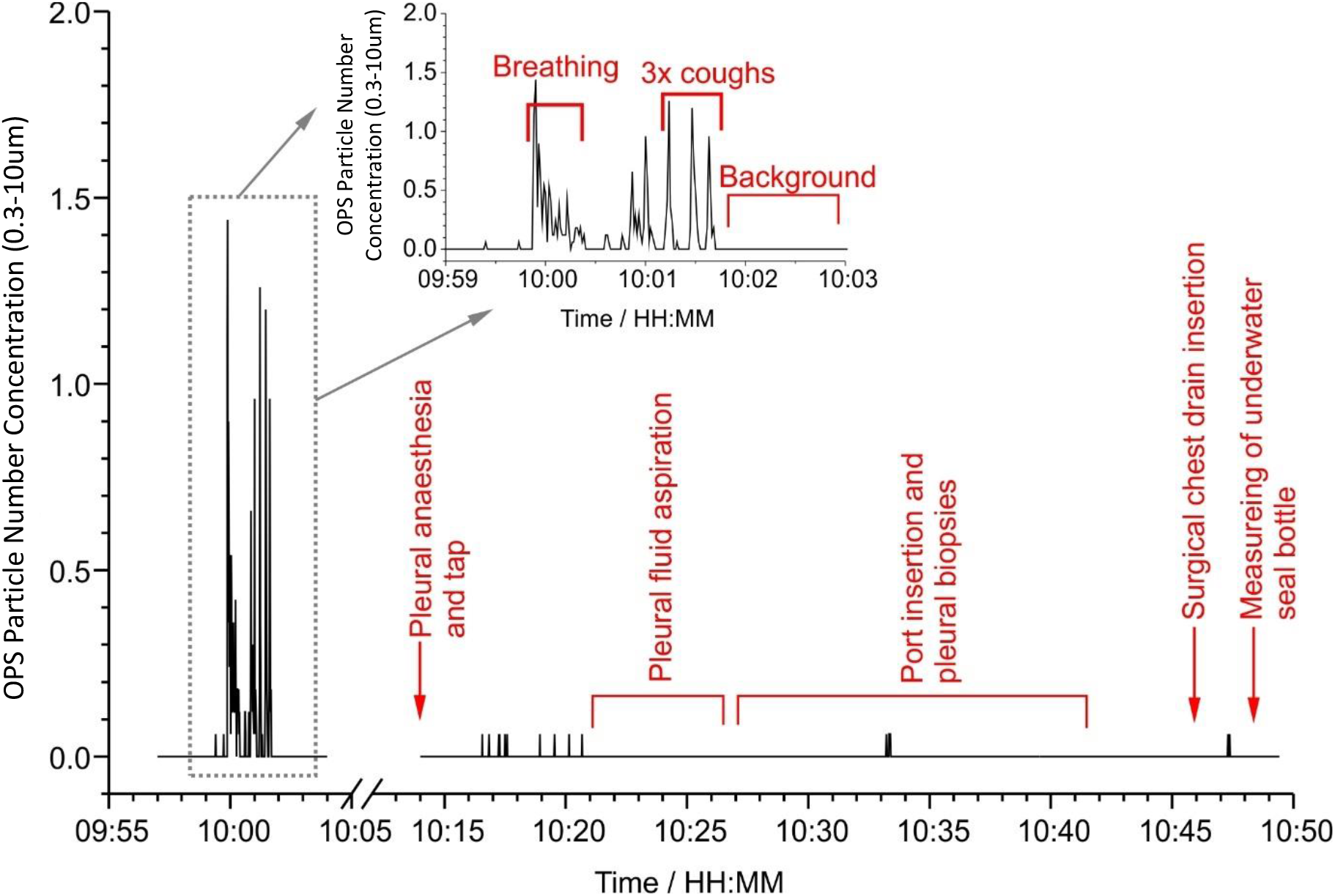
Particle concentration over time during a single thoracoscopy compared to aerosol production from the patient breathing and coughing pre-procedure.

## Discussion

This study shows that percutaneous instrumentation of the pleura does not result in significant aerosol generation. Total aerosol production during these procedures was significantly below aerosols produced by breathing or coughing.

Current British Thoracic Society (BTS) guidelines recommend that “closed pleural procedures such as pleural aspirations and chest drain insertion can be undertaken in Level 1 PPE (surgical mask and visor, as well as gown and gloves)” whereas “open procedures such as thoracoscopy and IPC insertion, where pleural fluid may splash, should still be considered AGP. Therefore, Level 2 PPE should be worn (FFP3 mask, long sleeved gown, gloves, eye protection)”. On the basis of this evidence, pleural procedures are not aerosol generating and additional PPE (above that indicated for routine patient care) is not required, although eye protection should be worn given the risk of splash^(4)^.

It is well recognised that pleural procedures, especially those that generate a negative intrathoracic pressure (e.g. therapeutic thoracentesis) can induce a cough in participants. We would therefore recommend the patient be asked to wear a surgical facemask, which has been shown to significantly reduce aerosol produced during cough (https://www.medrxiv.org/content/10.1101/2021.01.29.21250552v1).

Pleural fluid management systems such as underwater seal chest tube bottles have also been seen as a source of aerosol generation with several studies advocating the use of antiviral filters. Duffy and colleagues assessed aerosol generation by bubbling air at different rates through an underwater seal bottle^(4)^, sampling a maximum aerosol concentration of particles (within the same size range to those studied here, 0.3–10µm) during the bubbling process of ∼4100ft^-3^, caused by atomization of the water. This equates to a peak concentration of ∼0.14cm^-3^, which is similar to the peak concentrations that we observed during the fluctuations of sampled aerosol concentration during pleural procedures. We show that the peak number concentration sampled during the pleural procedures was similar to that sampled during the background measurement, orders of magnitude smaller than that sampled during a cough and was never greater than the mean concentration sampled during a period of quiet breathing. However, given our sample size for underwater seal bottles is small (n=3) and the mitigating factors are simple we feel guidance should still encourage the use of viral filters or Thopaz devices until further evidence is gathered, especially in pneumothoraces with high air leaks.

In summary, using two methodologies to measure aerosol emission with no background aerosol interference, this study has shown that percutaneous pleural procedures are non-aerosol generating. We hope this will inform future iterations of guidelines on the appropriate use of PPE when performing these procedures.

## Data Availability

No specific datasets have been generated for data sharing.

## Acknowledgements

We would also like to thank all the patients and volunteers who took part in the AERATOR study.

## Contributorship statement

DTA, FWH and NAM developed the study idea. FKAG, FWH, JWD, BRB and JPR developed the sampling design. DTA, SS, FWH, HW, AD, GN, NAM collected the primary aerosol data. FKAG, SS, BRB and JPR analysed the aerosol data. All authors were involved in the writing of the manuscript.

